# EEG correlates of confusional state after traumatic brain injury

**DOI:** 10.1101/2025.07.30.25332366

**Authors:** Angela Comanducci, Chiara-Camilla Derchi, Tiziana Atzori, Chiara Valota, Pietro Arcuri, Pietro Davide Trimarchi, Michele Angelo Colombo, Arturo Chieregato, Marcello Massimini, Jorge Navarro

## Abstract

Post-traumatic amnesia (PTA), recently conceptualized as part of the broader syndrome known as post-traumatic confusional state (PTCS), marks a critical phase of recovery following moderate-to-severe traumatic brain injury (TBI). Indeed, this state is characterized not only by anterograde memory impairment but also by disorientation, agitation, and attention deficits. Given the phenotypic overlap between PTA/PTCS and delirium—both marked by fluctuating cognitive and attentional disturbances—EEG represents a promising tool for elucidating shared pathophysiological mechanisms. While delirium is typically associated with diffuse EEG slowing and the presence of slow-wave activity (SWA), thought to reflect underlying global cortical disruption, it remains unclear whether PTCS exhibits similar EEG underpinnings.

In this prospective longitudinal study, we assessed dynamic EEG correlates of PTCS using the Grand Total EEG (GTE) score. We enrolled 42 consecutive TBI patients, classifying them at baseline into PTA/PTCS or emerged from PTA/PTCS, based on validated clinical assessments. Patients with PTA/PTCS exhibited significantly higher baseline GTE scores compared to TBI controls, reflecting severe EEG abnormalities characterized by diffuse slowing and disrupted rhythmic activity. Longitudinal follow-up revealed significant EEG improvements paralleling clinical recovery, confirming EEG’s sensitivity to dynamic clinical changes. Furthermore, the severity of EEG abnormalities significantly correlated with the duration of PTA/PTCS, underscoring EEG’s potential as an objective biomarker for monitoring recovery trajectories. Notably, these findings were independent of pharmacological confounders, as medication regimens were not significantly different across groups and timepoints.

Our results support a reconceptualization of PTA/PTCS as a functional encephalopathy, analogous to delirium, with EEG slowing reflecting widespread yet reversible cortical dysfunction. By capturing these transient yet clinically critical changes, clinical EEG— quantified via GTE scores—offers a novel tool for diagnosing PTA/PTCS, stratifying its severity, and objectively monitoring its evolution in acute and subacute rehabilitation settings.

## Introduction

Individuals who sustain a moderate-to-severe traumatic brain injury (TBI) may experience loss of consciousness followed by a period of confusion and amnesia that can vary in length from brief to prolonged, before regaining full awareness ^1,2^. This phase, traditionally known as ‘post-traumatic amnesia’ (PTA) ^2,3^, plays a pivotal role in determining recovery trajectories after TBI. Notably, the duration of PTA has been shown to be a more robust prognostic indicator compared to other clinical measures, e.g. the Glasgow Coma Scale (GCS) ^4–6^ with prolonged PTA strongly associated with worse long-term functional outcomes, reduced independence and diminished quality of life.

Recently, the term ‘post-traumatic confusional state’ (PTCS) ^2,7^ has been introduced to better capture the multifaceted phenomenology of this condition, typically observed after TBI. Patients in PTA/PTCS usually experience not only significant memory impairment but also disorientation and attention deficits, often accompanied by fluctuating levels of consciousness, agitation and sleep-wake disturbances.

The clinical manifestations of PTA/PTCS share substantial similarities with delirium, a functional encephalopathy characterized by an acute and fluctuating onset of confusion, along with disturbances in attention, cognition, and perception, that occurs without evidence of structural brain damage ^8^. Despite their distinct etiologies, this overlap raises important questions about the shared pathophysiological mechanisms underlying these conditions.

Electroencephalography (EEG) has proven to be an invaluable tool in elucidating brain dysfunction in encephalopathies, such as delirium, where it typically reveals a generalized slowing of background activity and the presence of diffuse slow-wave activity (SWA) ^9–11^.

To quantify the severity of encephalopathies across different etiologies, EEG-based measures such as the Grand Total EEG Score (GTE) ^12–14^, have been developed. This composite measure aggregates various EEG domains, including background activity and the presence of diffuse and focal SWA, into a single score.

Despite the clinical similarities between delirium and PTCS, it is unknown whether the two syndromes share common EEG abnormalities. Specifically, it has not yet been established whether EEG slowing could serve as a definitive marker of PTCS or how the presence of SWA may relate to the clinical course and the potential reversibility of this condition.

To address these questions, this study systematically investigates EEG abnormalities in individuals with PTA/PTCS following moderate-to-severe TBI. Specifically, we seek to determine whether EEG slowing, quantified using the GTE, is a core neurophysiological feature of PTCS and whether it relates to clinical recovery.

Specifically, we will compare in a cross-sectional design, patients with PTA/PTCS to those who had recovered from an early confusional phase and were no longer confused at the time of admission. Further, we will longitudinally track GTE changes in those patients recovering from PTA/PTCS during the rehabilitation stay, to characterize the neurophysiological trajectory of evolution of confusion. Finally, we will explore correlations between EEG slowing and key clinical variables, such as the severity and duration of PTA/PTCS.

## Material and methods

### Participants and study design

During the two-year study period (June 2021–June 2023), prospective participants were identified from consecutive admissions transitioning from the Intensive Care Unit (ICU) to the inpatient Acquired Brain Injury Intensive Rehabilitation Unit (IRU) at IRCCS Fondazione Don Carlo Gnocchi in Milan, Italy.

To minimize clinical heterogeneity and ensure a cohort with consistent injury timelines, severity, and underlying pathophysiology, inclusion criteria were defined as follows: (1) subacute TBI (within three months post-injury); (2) moderate to severe TBI, as PTA is a recognized hallmark of both severity levels; (3) presence of diffuse axonal injury confirmed by neuroimaging, as a key pathophysiological mechanism of TBI; (4) relatively rapid transition to PTA/PTCS from an acute disorder of consciousness (DoC) (defined as coma, unresponsive wakefulness syndrome/vegetative state (UWS/VS), or minimally conscious state (MCS) lasting less than 28 days, in line with current DoC guidelines ^15^; and (5) a diagnosis of PTA at ICU discharge, evidenced by a Level 4 score on the Levels of Cognitive Functioning Scale (LCF) ^16^ indicative of confused and agitated behavior.

Exclusion criteria included (1) severe deafness, aphasia or unstable clinical conditions; (2) a linguistic barrier that may hinder accurate assessment when conducting neuropsychological tests; (3) age below 18 or above 75; (4) insufficient medical documentation from ICU; (5) major focal lesions on neuroimaging (for example, subarachnoid hemorrhage, subdural hemorrhage, or focal ischemic damage); (6) a documented neurological or psychiatric disorder (particularly neurodegenerative or acquired conditions affecting cognitive domains, e.g. dementia); (7) medical conditions likely to influence clinical diagnosis or EEG (such as severe hepatic or renal insufficiency and a subcontinuous or abundant epileptiform activity detected on a standard EEG acquisition); (8) a history of decompressive craniectomy. These latter criteria (5-8) were specifically introduced to mitigate any potential confounding effects on EEG cortical activity.

The first EEG recording (T0) was scheduled within the first 7–10 days after admission to the IRU. During this period, clinically-stable patients who met the inclusion and exclusion criteria were recruited and repeatedly evaluated using the Galveston Orientation and Amnesia Test (GOAT)^17^ (every two days) and the Confusion Assessment Protocol (CAP) (twice weekly) ^17^ to assess their confusional state, classifying them as either in PTA/PTCS (still confused) or as TBI controls (having emerged from PTA/PTCS).

Patients still in PTA/PTCS at T0 continued to be monitored with the GOAT and the CAP to track recovery and document the evolution of PTA/PTCS. For these patients, a second EEG recording session (T1) was scheduled upon stable emergence from the confusional state or, if emergence did not occur, at discharge (but no later than two months post-admission, in accordance with local hospitalization regulations).

The informed consent was obtained from the patients themselves when they were capable; otherwise, it was acquired from their legal surrogates. This study was conducted following the principles of the Helsinki Declaration and was approved by the Ethics Committee of IRCCS Fondazione Don Carlo Gnocchi in Milan, Italy (ethics committee IRCCS Regione Lombardia, protocol number 32/2021/CE_FdG/FC/SA).

Details of the study design, including the timing of assessments and EEG recordings, are illustrated in Figure 1.

**Fig. 1.**
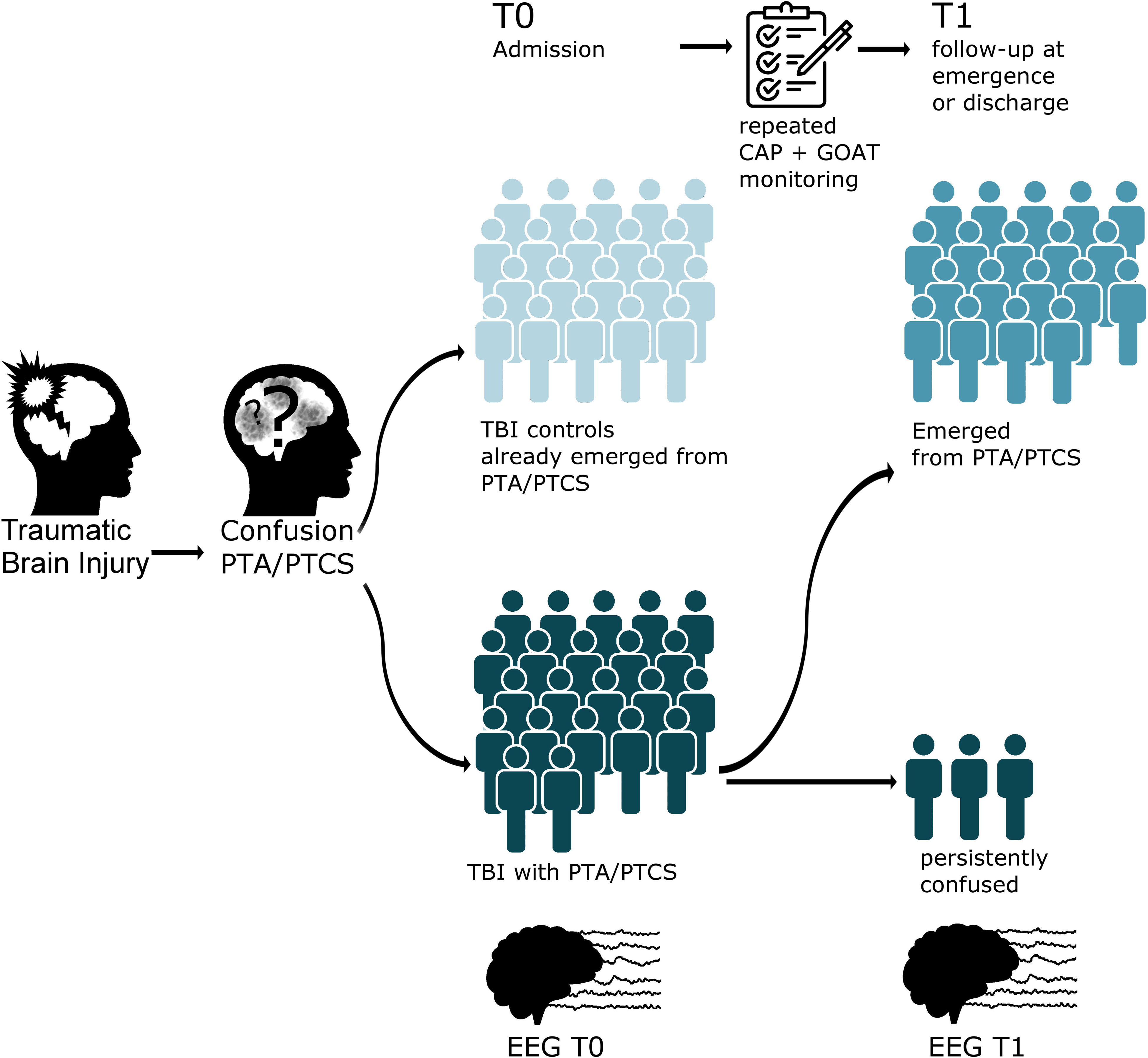
Diagram illustrating the study cohort and the timeline of EEG evaluations. After discharge from intensive care, all patients with Traumatic Brain Injury were in a state of PTA. Recruitment (T0) occurred in the intensive rehabilitation unit, where patients were assessed and classified into controls (light blue) and confused (PTA/PTCS, dark blue) using the CAP and the GOAT. For those classified as confused at T0, clinical monitoring initiated at T0 continued until the resolution of confusion. At the follow-up EEG recording (T1), these patients were further stratified into those who emerged from confusion (intermediate blue) and those who remained persistently confused (dark blue) upon discharge. PTA/PTCS: post-traumatic amnesia/post-traumatic confusional state; CAP: Confusion Assessment Protocol; GOAT: Galveston Orientation and Amnesia Test.

### PTA/PTCS assessment tools and clinical variables

To diagnose PTA/PTCS and assess its duration prospectively, we used two validated clinical scales, the GOAT and the CAP, which are widely recognized and commonly used across Europe ^17,7^.

The GOAT was designed to evaluate orientation to person, place, time, and event, providing a measure for determining the resolution of PTA ^18^. Scores range from 0 to 100, with higher scores indicating better orientation and memory function. PTA is considered present when the GOAT score is below 75. For this study, we employed the validated Italian version of the GOAT ^19^.

The CAP was developed to systematically measure seven multidomain key symptoms of PTCS: (1) cognitive impairment, (2) disorientation, (3) agitation, (4) symptom fluctuations, (5) nighttime sleep disturbance, (6) decreased daytime arousal, and (7) psychotic symptoms. Items are summed to calculate a total CAP score ranging from 0 (no symptoms of confusion) to 7 (all symptoms of confusion are present). PTCS was diagnosed in the presence of four factors, or three if one is disorientation. The CAP has demonstrated strong construct and criterion validity, aligning well with DSM-IV (Diagnostic and Statistical Manual - fourth edition) -based delirium diagnosis ^20,21^. It was developed using reference scales commonly used to assess PTA and delirium (e.g., GOAT, Agitated Behavior Scale, Delirium Rating Scale, 4AT). For our study, we employed the Italian translations of these scales, which have been validated ^19,22–24^. PTCS was also categorized as mild (CAP score <5) or severe (CAP score ≥5), using the cut-off of 5 as a practical threshold to identify clinically relevant cases, since a CAP score of 3 or 4 represents the minimum threshold for defining PTCS.

Given the rapidly fluctuating nature of PTA/PTCS, stable emergence from the confusional state was defined as two consecutive GOAT scores of 75 or higher ^25,26^ and two consecutive CAP assessments below the clinical threshold for clearing PTA ^27^.

For each patient, we collected several clinical variables, including demographic data (gender, age, and years of education) and injury-related variables such as the type of TBI, the maximal GCS score during the ICU stay, the time from TBI to the first EEG recording (T0), and the Disability Rating Scale (DRS) ^28^ score at IRU admission (Table 1).

**Table 1.** Demographic and clinical characteristics of patients with PTA/PTCS and without (TBI controls) confusional state. Continuous variables are reported as median and interquartile range, while categorical variables are expressed as frequencies and percentages. Comparisons for clinical and demographics data between clinical groups were performed using non-parametric tests, such as the Mann-Whitney U test for continuous variables and the chi-square test for categorical variables. NA: not available; CAP: Confusion Assessment Protocol; df: degrees of freedom; DRS: Disability Rating Scale; GOAT: Galveston Orientation and Amnesia Test; MVA: motor vehicle accident; PTA/PTCS: post-traumatic amnesia/post-traumatic confusional state; TBI: traumatic brain injury.

| Variable | PTA/PTCS (n=22) | TBI controls (n=20) | Test | Statistic | df | p-value |
| --- | --- | --- | --- | --- | --- | --- |
| Age | 38.4 [27.9-54.7] | 36.4 [28.2-53.6] | Mann-Whitney U | 228 | NA | 0,8502 |
| Education | 13.0 [12.2-13.0] | 13.0 [13.0-13.0] | Mann-Whitney U | 201 | NA | 0,5995 |
| GCS total score in ICU | 6.0 [3.2-7.8] | 8.0 [5.5-10.0] | Mann-Whitney U | 159,5 | NA | 0,1269 |
| Time from TBI to T0 (days) | 24.0 [16.2-33.5] | 24.0 [18.8-29.5] | Mann-Whitney U | 217 | NA | 0,9498 |
| DRS T0 | 18.5 [17.0-20.0] | 15.0 [13.0-18.0] | Mann-Whitney U | 341 | NA | 0,0021 |
| CAP total score T0 | 5.0 [4.2-6.0] | 0.0 [0.0-0.2] | Mann-Whitney U | 0 | NA | <0.0001 |
| GOAT T0 | 37.0 [27.0-45.0] | 100.0 [90.5-100.0] | Mann-Whitney U | 440 | NA | <0.0001 |
| duration PTA/PTCS | 68.5 [49.2-102.8] | 30.0 [23.8-42.0] | Mann-Whitney U | 377,5 | NA | 0,0001 |
| Gender | 19 male (86.4%) | 17 male (85.0%) | Chi-squared | 0 | 1 | 1 |
| Type of TBI | fall 4 (18.2%) | fall 4 (20.0%) | Chi-squared | 0,04 | 2 | 0,9811 |
|  | MVA 16 (72.7%) | MVA 14 (70.0%) |  |  |  |  |
|  | pedestrian 2 (9.1%) | pedestrian 2 (10.0%) |  |  |  |  |

Finally, to document any potential drug impact both on EEG and the evolution of the confusional state, we focused on three pharmacological classes widely used in TBI and known for their effects on the central nervous system: benzodiazepines, antipsychotics, and antiepileptics. For each patient, the total daily dose (expressed in mg/day) of each drug class was calculated over the 7-day period preceding the time-points of evaluation. Equivalent doses were calculated using established conversion methods: antipsychotic doses were expressed as chlorpromazine equivalents ^29^, benzodiazepine doses as diazepam equivalents ^30^, and antiepileptic doses as the cumulative percentage of the Defined Daily Dose (DDD) for each antiepileptic drug, following the recommendations of World Health Organization ^31^.

### EEG protocol, recording and analysis

Because of the transitory nature of confusional symptoms, which can fluctuate on the time-scale of days ^20,32^ EEG was performed only when subjects had a documented GOAT and CAP assessment on the same day.

EEG was recorded during a resting-state condition with eyes open and during a reactivity protocol, each lasting approximately 10 minutes. During the reactivity protocol, patients were instructed to open and close their eyes for at least 30 seconds at least three times, and to open and close their right and left hand for 30 seconds each.

To ensure optimal recording conditions and minimize psychomotor agitation, especially in confused patients, EEG acquisition was conducted in a quiet room outside the ward, free from any distracting stimuli.

Patients were seated in a comfortable chair and constantly supervised by one or, at most, two operators to promote a calm environment and facilitate patient compliance.

EEG recordings were conducted in the morning, a time when the patient was typically most alert, cooperative, and not yet fatigued from rehabilitative activities.

Signs of drowsiness were continuously monitored both through EEG activity and behavioral observation to ensure a sustained level of wakefulness throughout the recording session. Sessions were avoided if the patient had experienced a sleepless night or if daytime sleepiness was observed.

A 32-channel EEG amplifier (BrainAmp DC, Brain Products, Germany) was used to record EEG signals with electrodes placed on a cap according to the International 10/20 System. Electrode impedances were maintained below 5 kΩ, and the signals were acquired at a sampling rate of 1000 Hz. All electrodes were referenced to FCz with AFz serving as the ground ^33,34^. Additionally, two pairs of peripheral electrodes were placed: one for horizontal electrooculography (EOG) and one for electrocardiography (EKG).

EEGs were visually rated using the extended version of the GTE score ^35^, which integrates four main domains of EEG activity—rhythmic background activity, diffuse SWA, focal SWA, and epileptiform abnormalities—into a single score ranging from 0 to 54, providing a detailed evaluation of background activity and superimposed SWA dynamics.

Importantly, the scoring of background activity domain is not solely based on the dominant frequency but includes additional physiologically-relevant parameters such as spatial gradient, amplitude and symmetry. Similarly, in the SWA domain, factors like the presence of paroxysmal activity (e.g., frontal intermittent rhythmic delta activity, FIRDA), as well as the prevalence during the recording and the reactivity of SWA, are assessed. Scores for each item may range from 0 (normal) to 5 (suggestive of a severely abnormal encephalopathy), offering a biologically grounded framework for evaluating brain dysfunction and its evolution.

EEGs were visually analyzed and scored by two experts in clinical neurophysiology of brain-injured patients (AC and TA). Consensus scoring by the two reviewers on each GTE subcomponent, as well as on the total GTE score, was used to rate each EEG recording, with both reviewers blind to the clinical data during scoring.

For a detailed description of GTE domains, scoring criteria and associated items, refer to Supplementary Table 1.

### Statistical analysis

All statistical analyses were performed using R software (Version 4.2.2) ^36^. All collected variables were initially assessed for normality of distribution and homogeneity of variance using diagnostic tools, including histograms, Shapiro-Wilk tests and Levene’s test.

For cross-sectional comparisons of GTE scores between groups, Welch’s t-test was employed when variances were unequal, with effect sizes calculated using Cohen’s d. Longitudinal within-subject comparisons were conducted using the t-paired test, with effect sizes calculated using Cohen’s d. Correlations between PTA duration and GTE scores at emergence from PTA/PTCS were analyzed by means of Spearman’s rank correlation coefficient (ρ). Comparative analyses of equivalent daily drug doses across clinical groups and time points were conducted using non-parametric tests when normality assumptions were violated. Effect sizes were interpreted according to standard conventions (Cohen’s d: 0.2–0.5 small, 0.5–0.8 moderate, >0.8 large).

Detailed clinical data (CAP scores and subscales, including GOAT, in Supplementary Table 2; pharmacological treatments with equivalent doses in Supplementary Table 3) and EEG features (GTE scores and subscores for each domain in Supplementary Table 4) for each individual participant across time points are available in the Supplementary Section.

### Sample Size Calculation

The sample size for this study was calculated using G*Power software to ensure sufficient power to detect significant differences in EEG analysis between patients with and without PTA/PTCS. Given the scarcity of literature with EEG markers in PTA/PTCS, we referred to the only available study by Shah et al.^37^, which employed an EEG quantitative index (Delta-Alpha Ratio). This study reported an Area Under Curve of the Receiver Operating Curve (AUC) of 0.77, indicating a moderate discriminative ability. Utilizing the Cohen’s d formula transformed from AUC, an effect size of approximately 1.045 was derived, indicative of a large effect ^38^. To achieve a power of 80% and maintain an alpha level of 0.05, we determined that a minimum of 16 participants per group would be necessary. To account for potential dropouts and non-compliance, we decided to enroll at least 20 participants per group, increasing our total to at least 40 participants.

## Results

### 1. Study sample

As illustrated in Supplementary Figure 1 showing the Consolidated Standards Of Reporting Trials (CONSORT) flow diagram, we screened a total of 57 consecutive patients with TBI who met the inclusion criteria upon admission to our IRU. Of these, 13 were excluded due to one or more specific exclusion criteria (e.g., unstable clinical condition, age outside the predefined range, insufficient medical documentation, major focal lesion, previous neurodegenerative disorders, active epilepsy, or decompressive craniectomy). As a result, 44 participants were deemed eligible and written informed consent was obtained. One patient was later excluded from the study due to poor EEG recording quality with excessive artifacts. This resulted in a final sample of 42 eligible participants who were included and allocated into two groups: 22 participants in the PTA/PTCS group and 20 in the TBI control group.

Demographic and clinical characteristics of the participants in the PTA/PTCS and the TBI control group are summarized in Table 1. Both groups were predominantly male (PTA/PTCS: 19 male, 3 female; TBI control: 17 male, 3 female), and had similar median ages (38.4 vs. 36.4) and years of education (13.00). Mechanisms of injury were also comparable, involving mainly motor vehicle accidents (MVA) and falls. Maximal GCS scores reported in the ICU did not differ significantly between the groups (p=0.12). Although time from TBI to T0 was similar (median 24 days in both groups, p=0.95), the Disability Rating Scale (DRS) score at admission was significantly higher in PTA/PTCS (18.5) than in TBI controls (15.0, p=0.002). As expected, the CAP total score at T0 was markedly greater in patients with PTA/PTCS compared to not-confused patients (U=0, p<0.0001), whereas the GOAT score at T0 was lower (U=440, p<0.0001). Finally, the median duration of confusional state was also significantly longer at T0 in patients with PTA/PTCS (68.5 days) compared to TBI controls (30 days), indicating a more prolonged confusional state extending into the rehabilitation admittance period (U=377.5, p<0.0001).

### 2. EEG Patterns at baseline: comparison between TBI Control and PTA/PTCS and the relationship with severity

The results highlight the utility of the GTE score as an effective diagnostic marker for PTA/PTCS, capturing distinct neurophysiological profiles between confused and not-confused TBI patients. As shown in Figure 2, the distribution of GTE scores at baseline revealed significantly higher mean and standard deviation values in the PTA/PTCS group compared to TBI controls (Mean ± SD: 16.64 ± 4.50 vs. 5.10 ± 2.83, respectively). The plot reflects a substantially greater degree of EEG abnormalities in patients with PTA/PTCS, alongside a narrower range of variability observed in TBI controls. Statistical analysis supported the robustness of these findings, with Welch’s t-test confirming a highly significant difference between the two groups (t(35.75) = 10.04, p < 0.0001), with a very large effect size (Cohen’s d = 3.04).

**Fig. 2.**
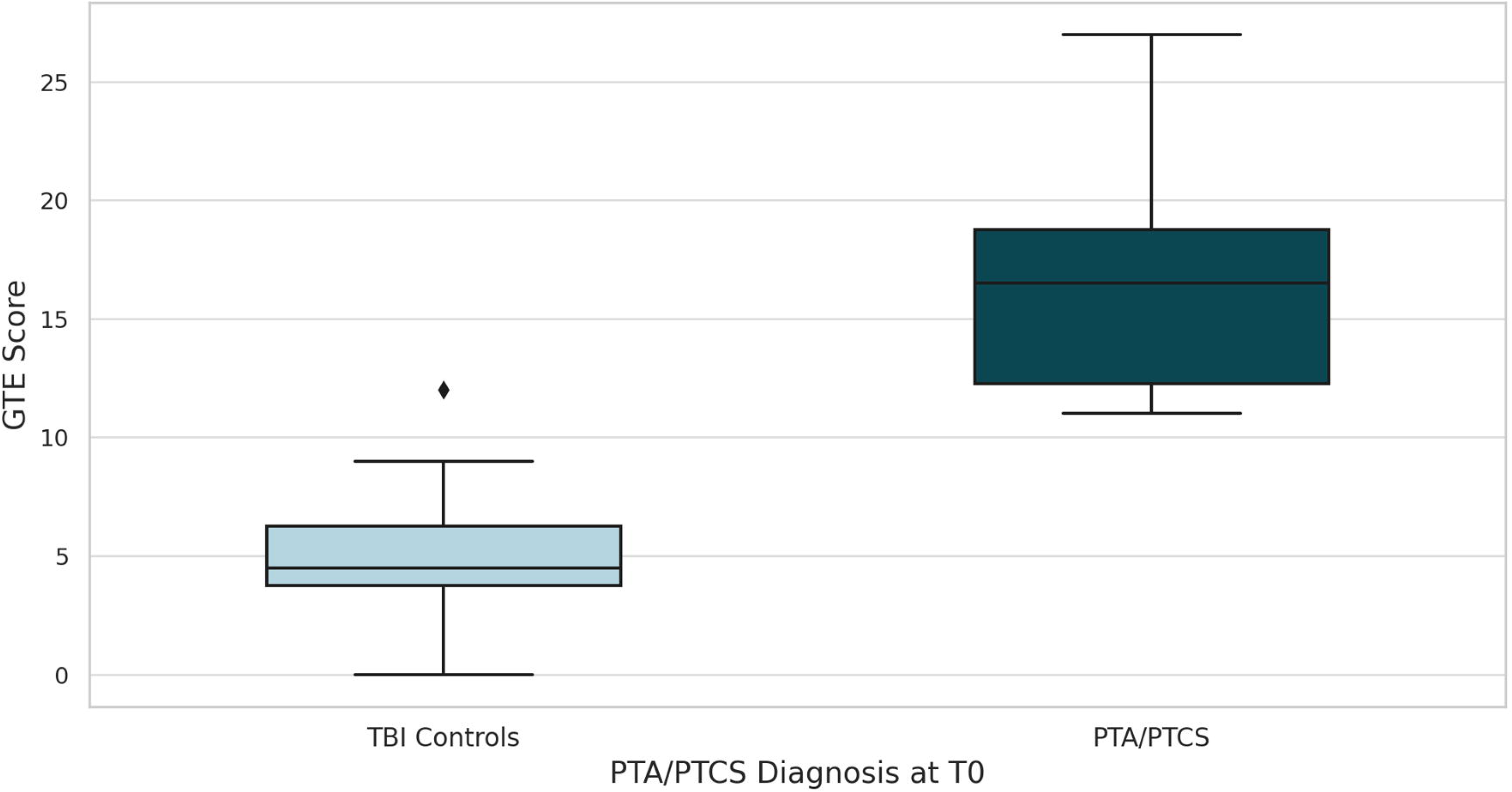
Cross-sectional comparison of GTE Score between TBI controls (light blue; left box-plot) and patients with PTA/PTCS (dark blue; right box-plot) at T0. GTE: Grand-Total EEG Score; PTA/PTCS: post-traumatic amnesia/post-traumatic confusional state, TBI: traumatic brain injury.

EEG proved to be a valuable tool for distinguishing between different levels of confusion severity, as determined by the CAP total score. As shown in Figure 3, patients with a CAP score <5, indicative of mild or absent PTCS, exhibited lower GTE scores (Mean ± SD: 7.12 ± 4.66) compared to those with a CAP score ≥5, indicative of severe PTCS (Mean ± SD: 17.69 ± 4.63). The histogram further illustrates this separation, showing that GTE scores for patients with mild PTCS are tightly clustered in a score range between 5 and 10, with a pronounced peak around 7. In contrast, patients with severe PTCS display a wider distribution of scores, ranging from 15 to 25, and peaking around 20, thus confirming a greater extent of EEG abnormalities. Statistical analysis using Welch’s t-test confirmed this significant difference (t(32.06) = 7.17, p < 0.0001) with a large effect size (Cohen’s d = 2.27).

**Fig. 3.**
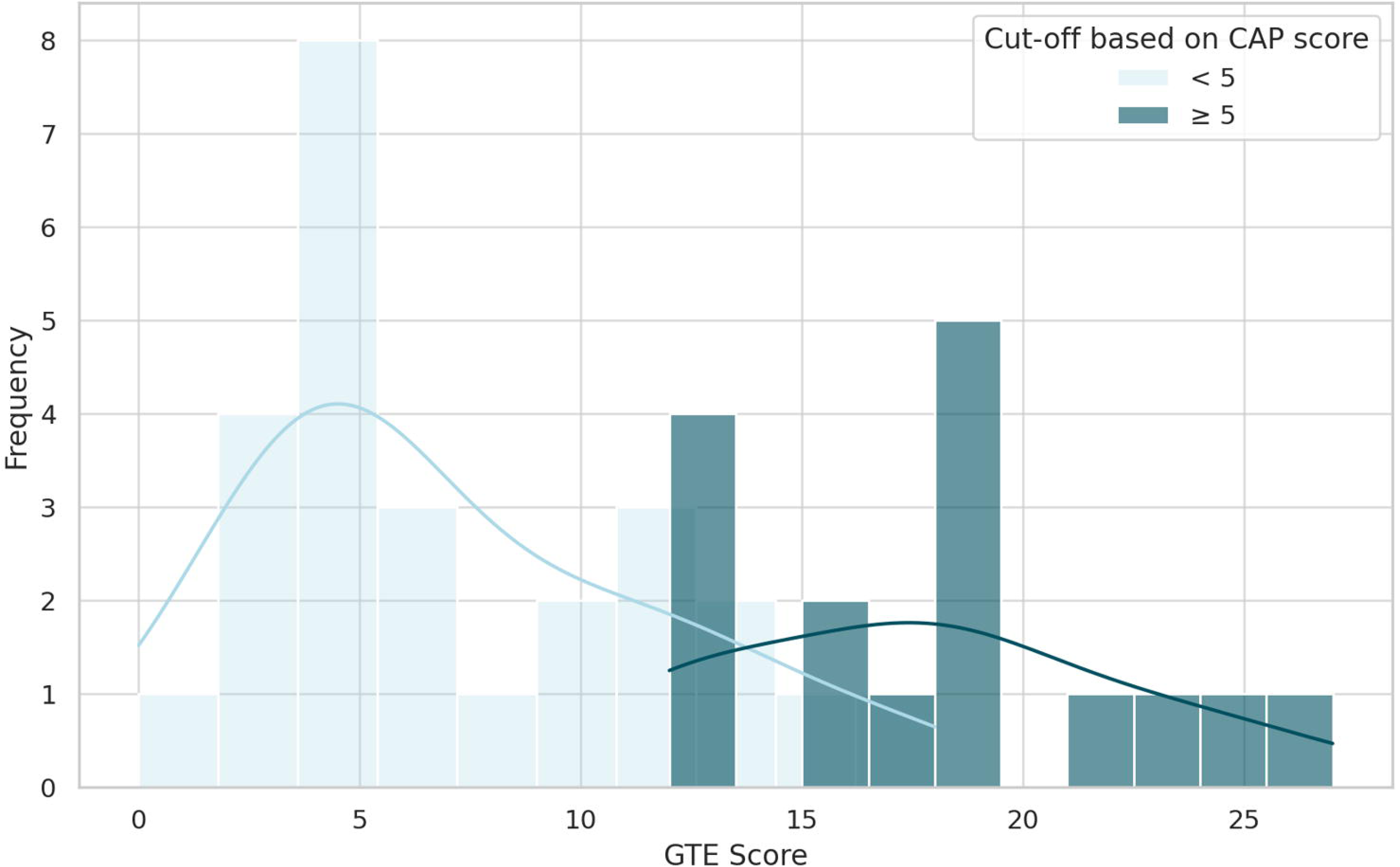
Relationship between CAP severity and GTE score (x-axis) at T0. The histogram illustrates the distribution of EEG-derived GTE scores according to a cut-off of 5 determined by CAP total scores (patients with CAP < 5 are depicted in light blue; dark blue is used for patients with CAP >= 5). The y-axis shows the frequency of observations for each GTE score. CAP: Confusion Assessment Protocol; GTE: Grand-Total EEG Score; PTA/PTCS: post-traumatic amnesia/post-traumatic confusional state.

### 3. Longitudinal EEG Dynamics during recovery from PTA/PTCS

Of the 22 participants diagnosed as confused at baseline, most (19/22; 86.4%) emerged from PTA/PTCS at follow-up (T1), with a median interval of 65 days from T0, while a few (3/22; 13.6%) remained persistently confused at discharge.

Longitudinal EEG evaluations effectively captured the evolution of clinical condition, demonstrating the utility of GTE score as an objective marker of neurophysiological improvement in PTA/PTCS. As illustrated in the individual trajectory plot (Fig. 4), a comparison between T0 and T1 revealed a clear reduction in GTE scores over time, reflecting neurophysiological recovery as patients emerged from PTA/PTCS. Overall, the median GTE score decreased from 16.5 (IQR: 6.5) at T0 to 8 (IQR: 3.75) at T1, marking a significant reduction in EEG abnormalities over time. This improvement was confirmed by a paired t-test which revealed a statistically significant difference between the two time points (t(21) = 8.03, p < 0.0001), with a very large effect size (Cohen’s d = 1.71). Most notably, all patients who recovered from PTA/PTCS from T0 to T1 showed some degree of improvement in their GTE score.

**Fig. 4.**
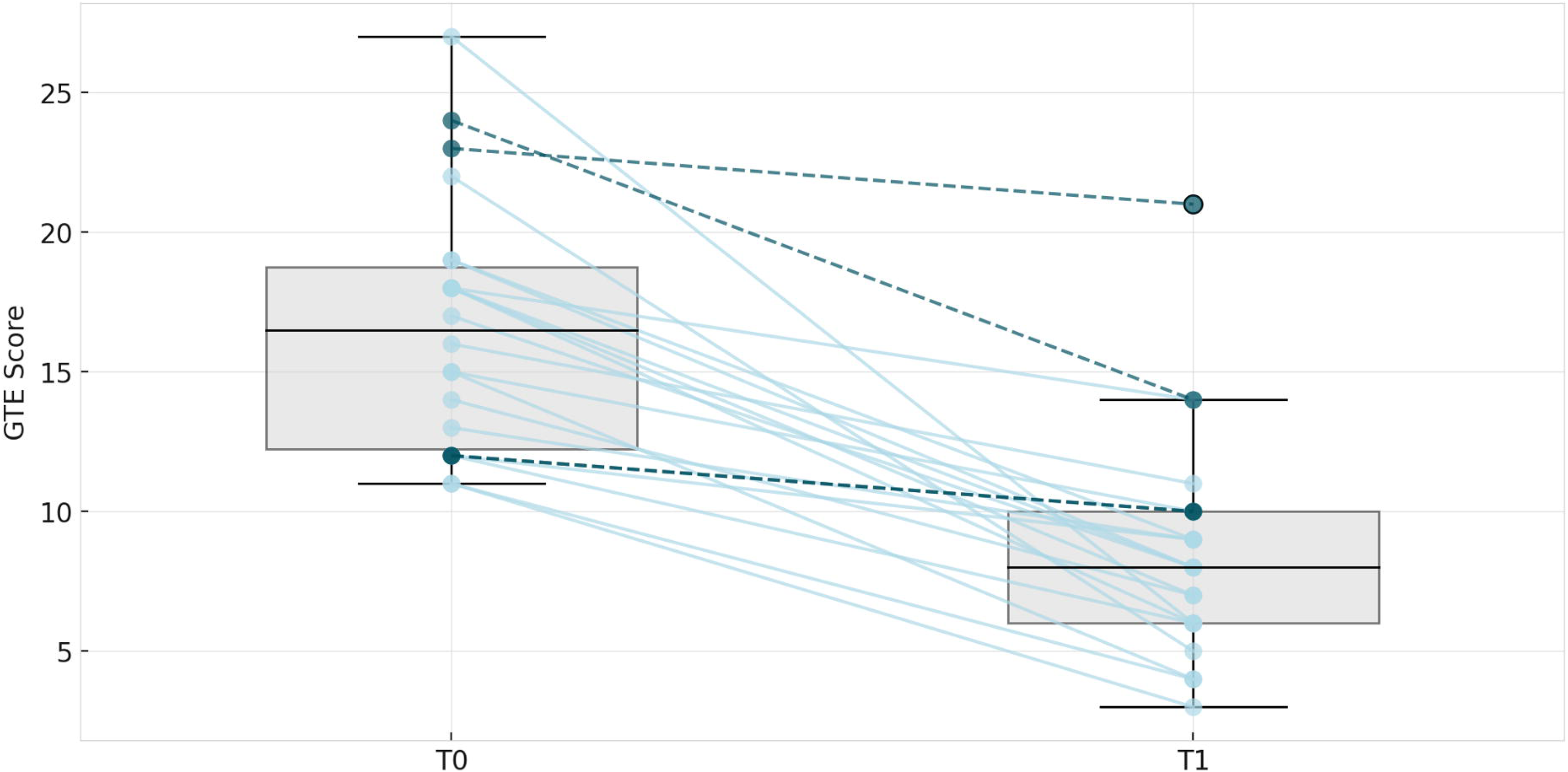
Individual trajectory plot showing the progression of GTE scores from time point T0 to T1 for patients with PTA/PTCS. The x-axis represents the time points of EEG recordings (T0 and T1), while the individual lines represent each patient’s score transition. All patients emerging from PTA/PTCS from T0 to T1 were represented by light blue lines. Patients highlighted with dark blue and dashed lines were the only three patients who remained persistently confused at follow-up. Box plots below the lines display the distribution of GTE scores at both time points, with medians marked in black, emphasizing the general trend in score changes over the observation period. GTE: Grand-Total EEG Score.

Despite this marked overall clinical and neurophysiological improvement, a subset of patients exhibited persistently elevated GTE scores at T1. Notably, all three patients who remained in persistent PTA/PTCS at follow-up had GTE scores ≥10 (10, 14, and 21), exceeding the 85th percentile of the patients emerging from PTA/PTCS at T1 (percentile threshold = 10). An exploratory Mann-Whitney U test confirmed that GTE scores at T1 were significantly higher in persistently confused vs. emerged patients (U = 58.0, p = 0.011), supporting the hypothesis that residual EEG abnormalities are associated with prolonged confusion.

### 4. Persistence and Correlates of EEG Abnormalities during PTA/PTCS Recovery

#### 4.1. Persistence of EEG Abnormalities in patients with a delayed Recovery from PTA/PTCS at T1

While the large majority of patients emerging from PTA/PTCS at T1 (19/22) exhibited a robust neurophysiological improvement, they continued to exhibit subtle EEG abnormalities even after recovery. Compared to TBI controls—patients who had already emerged from confusion at T0—those who recovered at T1 still displayed significantly higher GTE scores (Mean ± SD: 7.47 ± 2.65 vs. 5.1 ± 2.83; t(31.72) = 9.74, p < 0.0001, Cohen’s d = 3.15), indicative of a very large effect. As illustrated in the density plot (Fig. 5), the distribution of GTE scores further highlights this difference: patients who emerged from PTA/PTCS at T1 exhibited a right-shifted distribution compared to TBI controls, whose scores were more tightly clustered around lower values. This suggests that, even after clinical resolution, neurophysiological alterations persist in patients with delayed recovery, indicating incomplete normalization of EEG patterns.

**Fig. 5.**
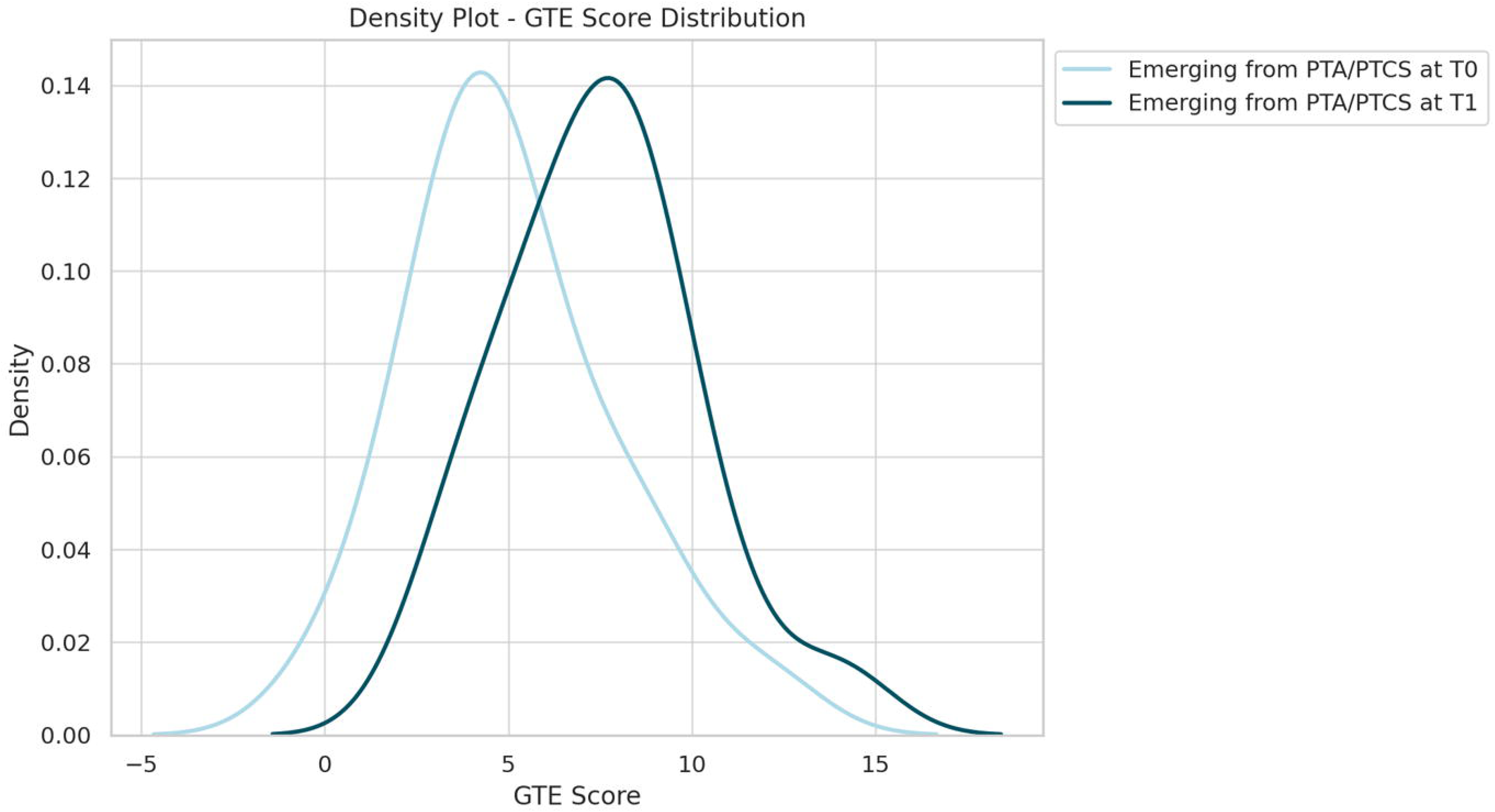
Kernel density plots of GTE scores comparing patients who emerged from PTA/PTCS at T0 (i.e. TBI controls; light blue) and those who were in PTA/PTCS at T0 and emerged only at T1 (dark blue). The distribution of GTE scores in patients emerging at T1 is broader and shifted toward higher values, indicating persistent neurophysiological alterations even after clinical recovery. In contrast, patients emerging at T0 exhibit a more compact distribution with lower scores, suggesting a more complete neurophysiological normalization. GTE: Grand-Total EEG Score; PTA/PTCS: post-traumatic amnesia/post-traumatic confusional state.

#### 4.2. Correlation between level of EEG impairment and PTA/PTCS duration

Longer durations of the confusional state were associated with more severe neurophysiological impairment, as demonstrated by the positive linear relationship between PTA/PTCS duration and GTE scores measured after symptom resolution in patients who successfully emerged from PTA/PTCS at either T0 (n=20) or T1 (n=19) (Fig.6). Spearman’s rank correlation analysis confirmed a statistically significant moderate-to-strong relationship (rho = 0.56, p<0.0001). These findings underscore the potential of EEG as an objective paraclinical tool for quantifying confusional state duration and aiding in the clinical assessment of recovery trajectories.

**Fig. 6.**
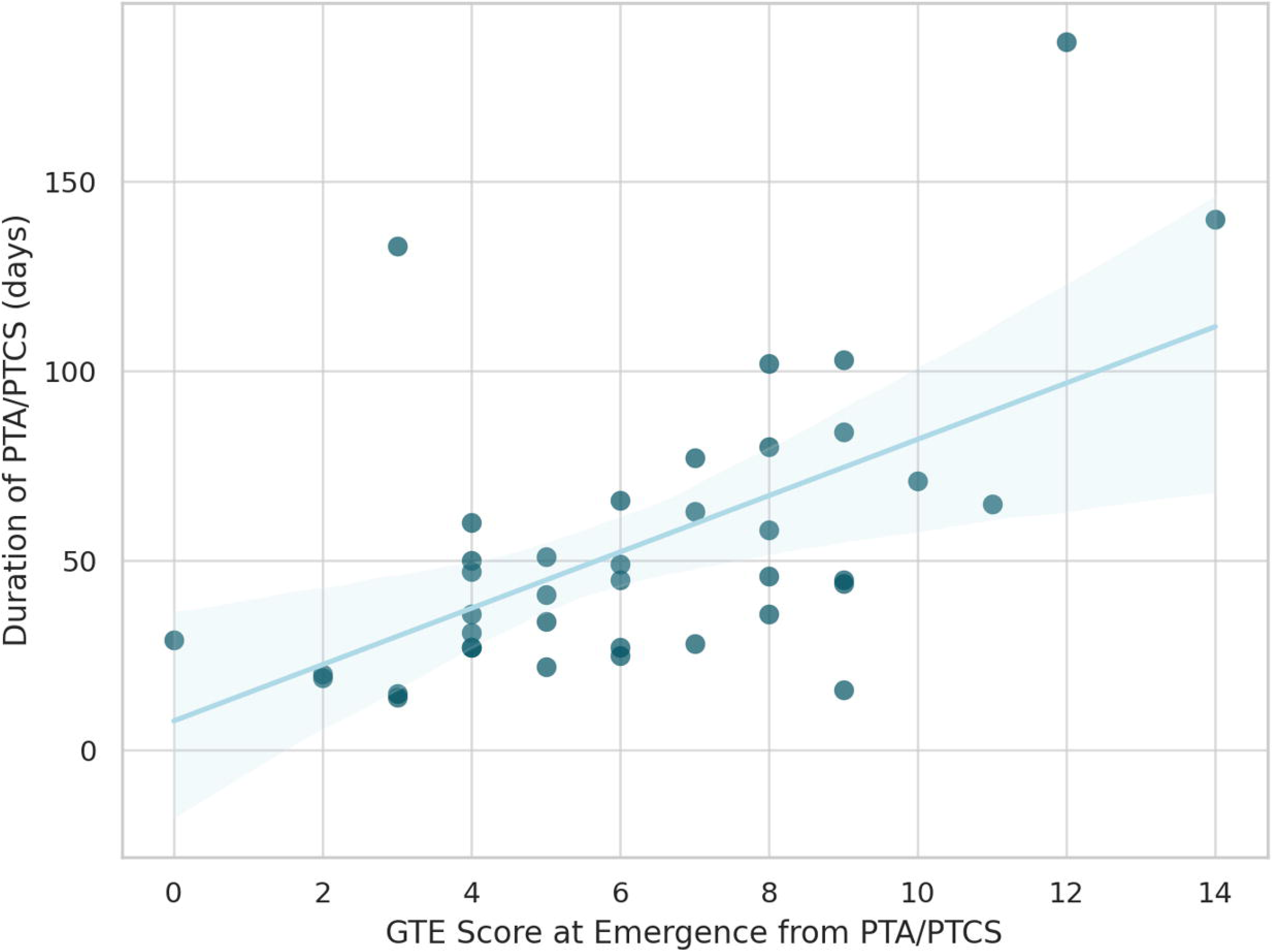
The scatter plot illustrates the linear relationship between the duration of PTA/PTCS and GTE score of patients at emergence from PTA/PTCS. The x-axis details the duration of PTA/PTCS in days, marking the period until the resolution of symptoms, while the y-axis displays the corresponding GTE scores measured at these specific time-points. Each point on the plot represents data from an individual patient emerged from PTA/PTCS at T0 or T1. A fitted regression line, with a 95% confidence interval shaded in light blue, is shown to delineate the trend. GTE: Grand-Total EEG Score; PTA/PTCS: post-traumatic amnesia/post-traumatic confusional state.

### 5. Stability of Pharmacological Regimens and their impact on the study

Differences in pharmacological treatment are unlikely to explain the neurophysiological and clinical differences observed between TBI controls and patients with PTA/PTCS at baseline or within the PTA/PTCS group longitudinally followed. Comparisons of sedative-hypnotic, antipsychotic, and antiepileptic drug use, expressed in equivalent doses and cumulative DDD, revealed no significant statistical differences either between the clinical groups at T0 or within the confused subsample over time (T0 vs. T1). Pharmacological regimens at single-patient level and the corresponding statistical comparisons are provided in Supplementary Tables 3 and 5.

## Discussion

This study provides novel insights into the neurophysiological mechanisms underlying PTA/PTCS and its recovery trajectories following moderate-to-severe TBI. By leveraging EEG-derived GTE score, we quantified the extent of brain dysfunction during PTA/PTCS and its resolution over time. At baseline, patients with PTA/PTCS exhibited pronounced EEG abnormalities compared to not-confused TBI controls, with minimal overlap between the two groups. EEG abnormalities partially normalized during follow-up in all patients who emerged from confusion; while patients who remained confused still exhibited pronounced abnormalities at follow-up. Moreover, the severity of EEG abnormalities after recovery was associated with the duration of the confusional state, suggesting that the extent of residual neurophysiological dysfunction reflects the clinical course.

### 1. Comparison with previous neurophysiologic literature in PTCS

The study by Shah and colleagues ^37^ represents the only available EEG investigation that focuses on PTCS, providing a basis for comparison with our findings. This study retrospectively explored the significance of delta-to-alpha frequency band power ratios from localized scalp areas in assessing the severity of the CAP total score and its correlation with long-term outcome. While increased delta activity and decreased posterior alpha rhythm have been already described in the acute phase of TBI, as well as in distinguishing mild TBI subjects from controls during eyes closed rest, most neurophysiological studies in TBI have not explicitly assessed confusional symptoms ^39–41^.

Our study extends beyond the analysis of a single spectral ratio by employing the GTE score, a comprehensive clinical EEG measure that evaluates multiple physiologically relevant domains of EEG activity. By leveraging the GTE score, we not only reinforce the importance of EEG markers in understanding PTCS but also enhance its diagnostic utility by explicitly comparing confused and not-confused patients. Our results strongly suggest that neurophysiological features, assessed with GTE, can not only accurately discriminate confused from not-confused patients, but that their longitudinal assessment can closely mirror recovery trajectories.

### 2. The GTE Score

The GTE score is a standardized EEG rating scale specifically designed to quantify the severity of encephalopathies. Originally developed for Alzheimer’s disease, it has been widely used and validated across a range of cognitive disorders due to its user-friendly nature and high reliability ^14,42,43^. Its applications extend to other encephalopathies characterized by cognitive dysfunction, including dementia with Lewy bodies, Parkinson’s disease, epilepsy, and autoimmune encephalitis ^35,42–45^.

Beyond its ease of use, the GTE score provides a biologically grounded and clinically interpretable method for objectively assessing brain dysfunction. Other clinical EEG classification systems ^46–48^ share these characteristics but were specifically designed for patients with acute (i.e. coma) or prolonged (i.e. UWS/VS and MCS) DoC, making them unsuitable for assessing awake conscious patients emerging from a subacute DoC.

Building on previous literature, our study offers several key advantages. First, we employed a longitudinal prospective design, capturing the evolution of PTCS during rehabilitation in the subacute phase, in addition to relying on cross-sectional retrospective data. Second, we show that the GTE was associated with the duration of the confusional state, further supporting its clinical relevance in tracking recovery.

### 3. PTCS as a Functional Encephalopathy and the Role of EEG slowing

This study is the first to apply the GTE score in PTCS, a transient and multifaceted cognitive disorder that shares key clinical features with delirium ^49–51^. Consistent with this clinical overlap, our findings reveal a neurophysiological parallel between PTCS and delirium ^11,52^, as evidenced by the combined presence of diffuse slowing and disruptions in rhythmic background activity, which emerged as hallmark EEG features of PTCS.

These results suggest shared pathophysiological mechanisms between the two conditions, reinforcing the role of EEG as a valuable tool for characterizing confusional states. Similar to delirium, where EEG slowing persists despite normalized arousal, this phenomenon in PTCS may reflect widespread cortical dysconnectivity rather than isolated arousal networks dysfunction. This supports the conceptualization of PTCS as a functional encephalopathy, characterized by reversible neurophysiological changes driven by impaired cortical-subcortical interactions following TBI.

The longitudinal reduction in GTE scores from baseline to T1 confirms the dynamic nature of PTA/PTCS, reflecting the progressive reorganization of brain networks during recovery and providing an objective temporal framework to track its progression.

### 4. Prognostic Implications of EEG in PTA/PTCS

While delirium has traditionally been viewed as part of a continuum leading to deeper states of unconsciousness such as stupor and coma ^53^, the modern concept of PTCS reframes it as a transitional state marking emergence from DoC following acquired brain injury ^54,55^.

The predominance of EEG slowing in this phase may represent both a marker of ongoing cortical network dysfunction and a reflection of the dynamic reorganization processes occurring during recovery. This distinction raises important questions about whether EEG slowing plays a causal role in maintaining the confusional state or acts as an epiphenomenon of widespread cortical dysfunction.

In delirium, generalized EEG slowing correlates with severity and is associated with worse clinical outcomes, prolonged hospitalization and increased mortality ^10,11,52,56^.

Understanding the prognostic role of EEG in PTA/PTCS is therefore crucial. Our preliminary findings suggest that patients with delayed emergence from PTA/PTCS exhibit incomplete normalization of EEG patterns, even after resolving their confusional state. This residual generalized slowing may reflect persistent cortical dysfunction and potentially predict poorer cognitive long-term outcome. Future studies should investigate whether specific GTE cutoffs could accurately predict recovery trajectories and guide clinical decision-making.

### 5. Limitations

#### Despite its strengths, this study has some limitations

First, since the duration of PTA is a well-established predictor of outcome, current guidelines, such as INCOG 2022 ^17^, recommend a daily assessment using validated tools like the Westmead Post-Traumatic Amnesia Scale (WPTAS) ^57^ until resolution. However, in this study, we did not include the WPTAS, opting instead for LCF, GOAT, and CAP, which are more commonly used in clinical practice across Europe ^17^. Future studies should consider integrating the WPTAS to explore its relationship with EEG findings and to determine PTA/PTCS resolution with greater temporal precision.

Second, while there is consensus on the need for validated tools to assess PTA/PTCS duration, no standardized instrumental criteria exist for quantifying its severity ^58^. To address this gap, we used the CAP total score, although it was not originally designed as a quantitative severity index. In this context, EEG emerges as a promising tool, providing an objective neurophysiological measure of brain dysfunction severity in PTA/PTCS.

However, the generalizability of our findings is limited. Multicentric studies with larger samples are needed to assess the intra- and inter-operator reliability of the GTE score and validate its use as a standardized EEG metric for diagnosing PTA/PTCS in subacute TBI patients.

Moreover, future research should complement visually assessed EEG features with relevant and interpretable quantitative metrics, such as the spectral exponent, a marker of broad-band slowing that showed robust diagnostic performance in the diagnosis of DoC ^33^. Integrating these approaches could offer deeper insights into the neurophysiological mechanisms underlying PTA/PTCS and enhance the accuracy of EEG-based assessments.

## Conclusions

In summary, our findings underscore the value of EEG in the assessment of confusional states and suggest that monitoring GTE scores can provide meaningful insights into recovery trajectories in patients with TBI. The reduction of EEG slowing in parallel with clinical improvement in PTA/PTCS may reflect a reversible, widespread cortical dysconnectivity rather than localized dysfunction of arousal networks. This supports the conceptualization of PTCS as a functional encephalopathy, with neurophysiological mechanisms partially overlapping with those of delirium. Overall, our results highlight the importance of integrating EEG into clinical practice to improve the understanding and management of confusional states in brain-injured patients.

### Transparency, Rigor, and Reproducibility

This study was not preregistered. Data analysis was conducted using SPSS. Data were collected up until June 2023. De-identified data from this study will be made available upon request, in accordance with IRB guidelines, by contacting the corresponding author.

## Data Availability

All data produced in the present study are available upon reasonable request to the authors

## Acknowledgements

We sincerely thank all the patients for dedicating their time to this study, even during the delicate period of rehabilitation.

## Authors Contributions (CRediT)

Angela Comanducci: Conceptualization (lead); Methodology (lead); Data acquisition and data analysis (lead); Writing – original draft (lead);

Chiara-Camilla Derchi: Methodology (lead); Data acquisition and data analysis (lead); Writing – original draft (lead);

Tiziana Atzori: Methodology (lead); Data acquisition (lead); Chiara Valota: Data acquisition (lead);

Pietro Arcuri: Data acquisition (lead);

Pietro Davide Trimarchi: Conceptualization (supporting); Methodology (supporting); Michele Angelo Colombo: Data analysis (supporting); Writing – review and editing (supporting);

Arturo Chieregato: Data acquisition (supporting);

Marcello Massimini: Conceptualization (supporting); Methodology (supporting); Writing – review and editing (supporting);

Jorge Navarro: Conceptualization (supporting); Methodology (supporting); Data acquisition (lead); Writing – review and editing (supporting).

## Declarations

Marcello Massimini is co-founder of Intrinsic Powers, a spin-off of the University of Milan. All the other authors have no competing interests to disclose.

## Ethics approval

This study was conducted following the principles of the Helsinki Declaration and was approved by the Ethics Committee of IRCCS Fondazione Don Carlo Gnocchi in Milan, Italy (ethics committee IRCCS Regione Lombardia, protocol number 32/2021/CE_FdG/FC/SA). The informed consent was obtained from the patients themselves when they were capable; otherwise, it was acquired from their legal surrogates.

