## Supplementary Table 1 for "EEG correlates of confusional state after traumatic brain injury"

|  | Rhythmic Background Activity | | | | | Diffuse Slow Wave Activity | | | | Focal Slow Wave Activity | | | | Epileptiform Abnormalities | |
| --- | --- | --- | --- | --- | --- | --- | --- | --- | --- | --- | --- | --- | --- | --- | --- |
| score | **frequency** | **reactivity** | **distribution of rhythmic BA** | **BA amplitude** | **asymmetry BA** | **diffuse SWA** | **paroxysmal activity** | **reactivity of diffuse SWA** | **prevalence of diffuse SWA** | **focal abnormalities** | **focal SWA** | **reactivity of focal SWA** | **prevalence of focal SWA** | **EA** | **location focal EA** |
| 0 | *>9* | *normal* | *occipital-temporal-parietal* | *>20 μV* | *symmetric* | *none* | *none* | *decreased with EO* | *none* | *none* | *none* | *decreased with EO* | *none* | *none* | *none* |
| 1 | *8-9* | *decreased with EO* | *up to central* | *<20 μV* | *asymmetric* | *intermittent theta* |  | *decreased with auditory but not EO* | *rare <1%* | *moderate unilateral* | *intermittent theta* | *decreased with auditory but not EO* | *rare <1%* |  | *moderate unilateral* |
| 2 | *7-8* | *absent with EO* | *up to frontal* |  |  | *intermittent theta + sporadic delta* |  | *decreased with pain only* | *occasional 1-9%* | *moderate bilateral* | *intermittent theta + sporadic delta* | *decreased with pain only* | *occasional 1-9%* | *sporadic* | *moderate bilateral* |
| 3 | *6-7* | *absent with somatosensory* |  |  |  | *intermittent theta + intermittent delta* | *paroxysmal SWA* | *no reactivity* | *frequent 10-49%* | *severe unilateral moderate contralateral* | *intermittent theta + intermittent delta* | *no reactivity* | *frequent 10-49%* | *frequent* | *severe unilateral moderate contralateral* |
| 4 | *4-6* | *absent with auditory* |  |  |  | *continuous theta + delta* |  |  | *abundant 50-89%* | *severe bilateral* |  |  | *abundant 50-89%* | *triphasic* | *severe bilateral* |
| 5 | *<4* | *absent* |  |  |  | *continuous delta* | *FIRDA* |  | *continuous >90%* | *multifocal* |  |  | *continuous >90%* | *PLEDs* | *multifocal* |

**Supplementary Table 1.** *Grand-Total EEG Score, sub-domains and items.*

*BA: background activity; EA: epileptiform abnormalities; EO: eyes opening; FIRDA: frontal intermittent rhythmic delta activity; PLEDs: periodic lateralized epileptiform discharges; SWA: slow wave activity.*
