## Supplementary Table 2 for "EEG correlates of confusional state after traumatic brain injury"

| condition | CAP total score T0 | CI T0 | GOAT T0 | ABS T0 | Fluctuations T0 | Sleep T0 | Arousal T0 | Psychotic symptoms T0 | CAP total score T1 | CI T1 | GOAT T1 | ABS T1 | Fluctuations T1 | Sleep T1 | Arousal T1 | Psychotic symptoms T1 |
| --- | --- | --- | --- | --- | --- | --- | --- | --- | --- | --- | --- | --- | --- | --- | --- | --- |
| TBI control 1 | 0 | 28 | 100 | 17 | 0 | 0 | 0 | 0 |  |  |  |  |  |  |  |  |
| TBI control 2 | 0 | 28 | 100 | 14 | 0 | 0 | 0 | 0 |  |  |  |  |  |  |  |  |
| TBI control 3 | 0 | 28 | 100 | 14 | 0 | 0 | 0 | 0 |  |  |  |  |  |  |  |  |
| TBI control 4 | 1 | 16 | 81 | 16 | 0 | 0 | 0 | 0 |  |  |  |  |  |  |  |  |
| TBI control 5 | 0 | 28 | 100 | 17 | 0 | 0 | 0 | 0 |  |  |  |  |  |  |  |  |
| TBI control 6 | 1 | 28 | 100 | 17 | 0 | 0 | 0 | 0 |  |  |  |  |  |  |  |  |
| TBI control 7 | 0 | 28 | 100 | 17 | 0 | 0 | 0 | 0 |  |  |  |  |  |  |  |  |
| TBI control 8 | 0 | 28 | 100 | 14 | 0 | 0 | 0 | 0 |  |  |  |  |  |  |  |  |
| TBI control 9 | 0 | 28 | 100 | 17 | 0 | 0 | 0 | 0 |  |  |  |  |  |  |  |  |
| TBI control 10 | 0 | 28 | 100 | 14 | 0 | 0 | 0 | 0 |  |  |  |  |  |  |  |  |
| TBI control 11 | 0 | 22 | 94 | 14 | 0 | 0 | 0 | 0 |  |  |  |  |  |  |  |  |
| TBI control 12 | 0 | 28 | 100 | 14 | 0 | 0 | 0 | 0 |  |  |  |  |  |  |  |  |
| TBI control 13 | 0 | 26 | 100 | 14 | 0 | 0 | 0 | 0 |  |  |  |  |  |  |  |  |
| TBI control 14 | 0 | 26 | 91 | 14 | 0 | 0 | 1 | 0 |  |  |  |  |  |  |  |  |
| TBI control 15 | 0 | 28 | 99 | 18 | 0 | 1 | 0 | 0 |  |  |  |  |  |  |  |  |
| TBI control 16 | 1 | 28 | 94 | 21 | 0 | 1 | 1 | 0 |  |  |  |  |  |  |  |  |
| TBI control 17 | 0 | 20 | 84 | 17 | 0 | 0 | 0 | 0 |  |  |  |  |  |  |  |  |
| TBI control 18 | 0 | 28 | 84 | 17 | 0 | 1 | 1 | 0 |  |  |  |  |  |  |  |  |
| TBI control 19 | 2 | 22 | 89 | 27 | 2 | 1 | 1 | 0 |  |  |  |  |  |  |  |  |
| TBI control 20 | 2 | 22 | 84 | 21 | 1 | 0 | 1 | 0 |  |  |  |  |  |  |  |  |
| PTCS 1 | 7 | 6 | 44 | 21 | 2 | 3 | 2 | 3 | 5 | 18 | 47 | 20 | 1 | 1 | 1 | 1 |
| PTCS 2 | 6 | 16 | 37 | 22 | 1 | 1 | 2 | 2 | 2 | 16 | 85 | 18 | 0 | 1 | 0 | 1 |
| PTCS 3 | 4 | 4 | 54 | 21 | 0 | 0 | 1 | 3 | 2 | 16 | 74 | 17 | 0 | 1 | 0 | 1 |
| PTCS 4 | 5 | 18 | 27 | 21 | 2 | 1 | 1 | 3 | 1 | 22 | 99 | 18 | 0 | 0 | 0 | 0 |
| PTCS 5 | 5 | 8 | 26 | 18 | 1 | 0 | 0 | 3 | 1 | 28 | 78 | 14 | 0 | 0 | 1 | 3 |
| PTCS 6 | 6 | 18 | 31 | 24 | 1 | 1 | 2 | 3 | 1 | 24 | 80 | 20 | 0 | 0 | 1 | 0 |
| PTCS 7 | 6 | 10 | 15 | 38 | 1 | 3 | 1 | 3 | 2 | 28 | 100 | 19 | 1 | 1 | 1 | 1 |
| PTCS 8 | 5 | 8 | 26 | 20 | 1 | 1 | 2 | 1 | 2 | 20 | 74 | 16 | 1 | 0 | 1 | 1 |
| PTCS 9 | 4 | 6 | 62 | 18 | 0 | 0 | 0 | 2 | 1 | 16 | 89 | 14 | 0 | 0 | 0 | 0 |
| PTCS 10 | 5 | 14 | 35 | 22 | 0 | 2 | 1 | 2 | 3 | 16 | 94 | 20 | 1 | 0 | 0 | 0 |
| PTCS 11 | 4 | 8 | 45 | 21 | 1 | 0 | 0 | 0 | 1 | 18 | 77 | 15 | 0 | 0 | 0 | 0 |
| PTCS 12 | 4 | 20 | 67 | 20 | 1 | 1 | 1 | 0 | 1 | 24 | 80 | 18 | 0 | 0 | 0 | 0 |
| PTCS 13 | 5 | 4 | 27 | 27 | 1 | 2 | 1 | 2 | 1 | 22 | 90 | 18 | 0 | 0 | 0 | 0 |
| PTCS 14 | 4 | 14 | 43 | 21 | 1 | 0 | 0 | 0 | 1 | 28 | 71 | 16 | 0 | 0 | 0 | 0 |
| PTCS 15 | 6 | 2 | 7 | 25 | 1 | 2 | 2 | 2 | 1 | 20 | 100 | 19 | 0 | 0 | 0 | 0 |
| PTCS 16 | 3 | 24 | 40 | 20 | 1 | 1 | 1 | 0 | 1 | 26 | 85 | 14 | 0 | 0 | 0 | 0 |
| PTCS 17 | 5 | 28 | 58 | 24 | 1 | 2 | 1 | 1 | 1 | 28 | 100 | 14 | 0 | 0 | 0 | 0 |
| PTCS 18 | 6 | 2 | 37 | 32 | 1 | 2 | 1 | 5 | 2 | 18 | 94 | 18 | 0 | 0 | 0 | 3 |
| PTCS 19 | 6 | 2 | 4 | 28 | 0 | 2 | 1 | 6 | 6 | 2 | 1 | 25 | 1 | 1 | 1 | 3 |
| PTCS 20 | 6 | 10 | 45 | 23 | 1 | 2 | 1 | 2 | 0 | 22 | 95 | 16 | 0 | 1 | 0 | 0 |
| PTCS 21 | 6 | 8 | 29 | 28 | 1 | 2 | 0 | 5 | 6 | 2 | 26 | 29 | 1 | 2 | 0 | 2 |
| PTCS 22 | 5 | 12 | 72 | 20 | 1 | 2 | 1 | 0 | 0 | 20 | 95 | 16 | 0 | 0 | 0 | 0 |

**Supplementary Table 2.** *Confusion Assessment Protocol (CAP) total scores and subscores for each patient across time points (T0 and T1).*

*ABS: agitated behavior scale; CI: cognitive impairment; GOAT: Galveston Orientation and Amnesia Test; PTCS: post-traumatic confusional state; TBI: traumatic brain injury.*
