## Supplementary Table 3 for "EEG correlates of confusional state after traumatic brain injury"

| condition | timepoint | Sedatives-Hypnotics (Diazepam equivalent, mg) | Antipsychotics (Chlorpromazine equivalent, mg) | Antiepileptics (Total Sum % Defined Daily Dose) |
| --- | --- | --- | --- | --- |
| TBI control 1 | T0 | 0 | 33 | 0 |
| TBI control 2 | T0 | 0 | 133 | 0 |
| TBI control 3 | T0 | 0 | 133 | 0 |
| TBI control 4 | T0 | 10 | 230 | 0 |
| TBI control 5 | T0 | 10 | 67 | 83 |
| TBI control 6 | T0 | 0 | 0 | 0 |
| TBI control 7 | T0 | 0 | 0 | 0 |
| TBI control 8 | T0 | 0 | 0 | 0 |
| TBI control 9 | T0 | 4 | 200 | 0 |
| TBI control 10 | T0 | 0 | 0 | 0 |
| TBI control 11 | T0 | 0 | 0 | 0 |
| TBI control 12 | T0 | 10 | 33 | 67 |
| TBI control 13 | T0 | 0 | 366 | 42 |
| TBI control 14 | T0 | 7 | 466 | 0 |
| TBI control 15 | T0 | 0 | 0 | 0 |
| TBI control 16 | T0 | 0 | 0 | 0 |
| TBI control 17 | T0 | 0 | 67 | 33 |
| TBI control 18 | T0 | 8 | 432 | 0 |
| TBI control 19 | T0 | 3 | 50 | 0 |
| TBI control 20 | T0 | 0 | 67 | 77 |
| PTCS 1 | T0 | 0 | 0 | 0 |
| PTCS 2 | T0 | 4 | 346 | 0 |
| PTCS 3 | T0 | 5 | 67 | 8 |
| PTCS 4 | T0 | 6 | 133 | 0 |
| PTCS 5 | T0 | 20 | 133 | 53 |
| PTCS 6 | T0 | 0.9 | 0 | 0 |
| PTCS 7 | T0 | 10 | 50 | 40 |
| PTCS 8 | T0 | 0 | 700 | 0 |
| PTCS 9 | T0 | 0 | 0 | 0 |
| PTCS 10 | T0 | 0 | 0 | 0 |
| PTCS 11 | T0 | 0 | 0 | 0 |
| PTCS 12 | T0 | 1 | 0 | 0 |
| PTCS 13 | T0 | 5 | 200 | 0 |
| PTCS 14 | T0 | 0 | 33 | 0 |
| PTCS 15 | T0 | 0 | 0 | 0 |
| PTCS 16 | T0 | 0 | 0 | 67 |
| PTCS 17 | T0 | 20 | 200 | 67 |
| PTCS 18 | T0 | 20 | 0 | 0 |
| PTCS 19 | T0 | 0 | 113 | 0 |
| PTCS 20 | T0 | 0 | 50 | 40 |
| PTCS 21 | T0 | 10 | 100 | 60 |
| PTCS 22 | T0 | 0 | 133 | 27 |
| PTCS 1 | T1 | 18 | 70 | 67 |
| PTCS 2 | T1 | 23 | 0 | 0 |
| PTCS 3 | T1 | 5 | 0 | 8 |
| PTCS 4 | T1 | 6 | 33 | 73 |
| PTCS 5 | T1 | 20 | 67 | 27 |
| PTCS 6 | T1 | 1.6 | 33 | 0 |
| PTCS 7 | T1 | 0 | 133 | 55 |
| PTCS 8 | T1 | 0 | 133 | 0 |
| PTCS 9 | T1 | 0 | 0 | 0 |
| PTCS 10 | T1 | 0 | 0 | 0 |
| PTCS 11 | T1 | 0 | 0 | 0 |
| PTCS 12 | T1 | 0 | 0 | 0 |
| PTCS 13 | T1 | 3 | ~166 | 47 |
| PTCS 14 | T1 | 0 | 33 | 0 |
| PTCS 15 | T1 | 5 | 0 | 0 |
| PTCS 16 | T1 | 0 | 0 | 67 |
| PTCS 17 | T1 | 20 | 200 | 47 |
| PTCS 18 | T1 | 5 | 50 | 0 |
| PTCS 19 | T1 | 10 | 2500 | 13 |
| PTCS 20 | T1 | 0 | 50 | 50 |
| PTCS 21 | T1 | 0 | 1500 | 7 |
| PTCS 22 | T1 | 0 | 166 | 67 |

**Supplementary Table 3.** *The main three pharmacological classes for each patient across time points (T0 and T1).*

*PTCS: post-traumatic confusional state; TBI: traumatic brain injury.*
