## Supplementary Table 4 for "EEG correlates of confusional state after traumatic brain injury"

| condition | timepoint | Rhythmic Background Activity | | | | | Diffuse Slow Wave Activity | | | | Focal Slow Wave Activity | | | | Epileptiform Abnormalities | | GTE total score |
| --- | --- | --- | --- | --- | --- | --- | --- | --- | --- | --- | --- | --- | --- | --- | --- | --- | --- |
|  |  | **frequency** | **reactivity** | **distribution of rhythmic BA** | **BA amplitude** | **asymmetry BA** | **diffuse SWA** | **paroxysmal activity** | **reactivity of diffuse SWA** | **prevalence of diffuse SWA** | **focal abnormalities** | **focal SWA** | **reactivity of focal SWA** | **prevalence of focal SWA** | **EA** | **location focal EA** |  |
| TBI control 1 | T0 | 1 | 0 | 0 | 0 | 0 | 0 | 0 | 0 | 0 | 1 | 1 | 0 | 1 | 0 | 0 | 4 |
| TBI control 2 | T0 | 0 | 0 | 0 | 0 | 0 | 2 | 0 | 0 | 1 | 0 | 0 | 0 | 0 | 0 | 0 | 3 |
| TBI control 3 | T0 | 0 | 0 | 0 | 0 | 0 | 1 | 0 | 0 | 1 | 1 | 1 | 0 | 2 | 0 | 0 | 6 |
| TBI control 4 | T0 | 1 | 0 | 0 | 1 | 0 | 2 | 0 | 0 | 1 | 0 | 0 | 0 | 0 | 0 | 0 | 5 |
| TBI control 5 | T0 | 0 | 1 | 1 | 0 | 0 | 0 | 0 | 0 | 0 | 0 | 0 | 0 | 0 | 0 | 0 | 2 |
| TBI control 6 | T0 | 1 | 1 | 0 | 0 | 0 | 0 | 0 | 0 | 0 | 0 | 0 | 0 | 0 | 0 | 0 | 2 |
| TBI control 7 | T0 | 1 | 0 | 0 | 0 | 0 | 2 | 3 | 0 | 2 | 0 | 0 | 0 | 0 | 0 | 0 | 8 |
| TBI control 8 | T0 | 1 | 1 | 1 | 0 | 0 | 2 | 0 | 0 | 2 | 1 | 2 | 0 | 2 | 0 | 0 | 12 |
| TBI control 9 | T0 | 1 | 1 | 0 | 0 | 1 | 1 | 0 | 0 | 1 | 0 | 0 | 0 | 0 | 0 | 0 | 5 |
| TBI control 10 | T0 | 0 | 1 | 0 | 0 | 0 | 1 | 0 | 0 | 2 | 0 | 0 | 0 | 0 | 0 | 0 | 4 |
| TBI control 11 | T0 | 0 | 0 | 0 | 0 | 0 | 0 | 0 | 0 | 0 | 0 | 0 | 0 | 0 | 0 | 0 | 0 |
| TBI control 12 | T0 | 0 | 0 | 0 | 0 | 0 | 1 | 0 | 0 | 2 | 0 | 0 | 0 | 0 | 0 | 0 | 3 |
| TBI control 13 | T0 | 0 | 0 | 0 | 0 | 0 | 0 | 0 | 0 | 0 | 1 | 1 | 0 | 2 | 0 | 0 | 4 |
| TBI control 14 | T0 | 0 | 0 | 0 | 0 | 1 | 0 | 0 | 0 | 0 | 1 | 1 | 0 | 2 | 0 | 0 | 5 |
| TBI control 15 | T0 | 0 | 0 | 0 | 0 | 0 | 0 | 0 | 0 | 0 | 1 | 1 | 0 | 2 | 0 | 0 | 4 |
| TBI control 16 | T0 | 0 | 0 | 0 | 0 | 0 | 0 | 0 | 0 | 0 | 1 | 1 | 0 | 2 | 0 | 0 | 4 |
| TBI control 17 | T0 | 1 | 0 | 1 | 0 | 0 | 2 | 0 | 0 | 2 | 0 | 0 | 0 | 0 | 0 | 0 | 6 |
| TBI control 18 | T0 | 0 | 0 | 1 | 0 | 0 | 2 | 0 | 0 | 2 | 1 | 2 | 0 | 1 | 0 | 0 | 9 |
| TBI control 19 | T0 | 1 | 1 | 2 | 0 | 0 | 1 | 0 | 0 | 2 | 0 | 0 | 0 | 0 | 0 | 0 | 7 |
| TBI control 20 | T0 | 2 | 0 | 1 | 0 | 0 | 2 | 0 | 0 | 1 | 1 | 2 | 0 | 0 | 0 | 0 | 9 |
| PTCS 1 | T0 | 3 | 2 | 1 | 0 | 0 | 3 | 3 | 1 | 4 | 2 | 2 | 1 | 2 | 0 | 0 | 24 |
| PTCS 2 | T0 | 2 | 2 | 1 | 0 | 1 | 2 | 0 | 0 | 2 | 3 | 3 | 0 | 2 | 0 | 0 | 18 |
| PTCS 3 | T0 | 0 | 1 | 1 | 0 | 1 | 1 | 0 | 0 | 1 | 1 | 1 | 0 | 1 | 2 | 1 | 11 |
| PTCS 4 | T0 | 2 | 1 | 1 | 0 | 0 | 3 | 3 | 0 | 2 | 1 | 1 | 0 | 1 | 0 | 0 | 15 |
| PTCS 5 | T0 | 2 | 2 | 1 | 0 | 0 | 3 | 5 | 0 | 4 | 0 | 0 | 0 | 0 | 0 | 0 | 17 |
| PTCS 6 | T0 | 0 | 1 | 1 | 0 | 0 | 3 | 3 | 1 | 3 | 1 | 2 | 1 | 3 | 0 | 0 | 19 |
| PTCS 7 | T0 | 3 | 2 | 2 | 0 | 0 | 3 | 0 | 0 | 2 | 0 | 0 | 0 | 0 | 0 | 0 | 12 |
| PTCS 8 | T0 | 2 | 2 | 2 | 0 | 0 | 3 | 5 | 1 | 3 | 0 | 0 | 0 | 0 | 0 | 0 | 18 |
| PTCS 9 | T0 | 2 | 1 | 1 | 0 | 1 | 3 | 0 | 0 | 3 | 2 | 1 | 0 | 2 | 0 | 0 | 16 |
| PTCS 10 | T0 | 1 | 1 | 1 | 0 | 1 | 2 | 0 | 0 | 2 | 2 | 2 | 0 | 3 | 0 | 0 | 15 |
| PTCS 11 | T0 | 1 | 2 | 1 | 1 | 1 | 2 | 0 | 0 | 2 | 1 | 2 | 0 | 2 | 2 | 1 | 18 |
| PTCS 12 | T0 | 0 | 1 | 1 | 1 | 1 | 2 | 0 | 0 | 1 | 1 | 2 | 0 | 2 | 0 | 1 | 13 |
| PTCS 13 | T0 | 2 | 2 | 2 | 0 | 0 | 3 | 0 | 0 | 3 | 0 | 0 | 0 | 0 | 0 | 0 | 12 |
| PTCS 14 | T0 | 0 | 1 | 2 | 0 | 1 | 2 | 0 | 0 | 1 | 1 | 1 | 0 | 2 | 0 | 0 | 11 |
| PTCS 15 | T0 | 2 | 2 | 2 | 0 | 1 | 3 | 3 | 1 | 3 | 3 | 3 | 0 | 4 | 0 | 0 | 27 |
| PTCS 16 | T0 | 1 | 1 | 1 | 0 | 0 | 2 | 5 | 0 | 2 | 1 | 1 | 0 | 0 | 0 | 0 | 14 |
| PTCS 17 | T0 | 1 | 1 | 1 | 0 | 0 | 3 | 3 | 1 | 3 | 1 | 2 | 0 | 2 | 0 | 0 | 18 |
| PTCS 18 | T0 | 1 | 2 | 1 | 0 | 0 | 3 | 3 | 2 | 3 | 1 | 1 | 0 | 2 | 0 | 0 | 19 |
| PTCS 19 | T0 | 2 | 2 | 2 | 0 | 0 | 3 | 0 | 1 | 2 | 0 | 0 | 0 | 0 | 0 | 0 | 12 |
| PTCS 20 | T0 | 3 | 1 | 1 | 1 | 1 | 2 | 0 | 1 | 2 | 0 | 0 | 0 | 0 | 0 | 0 | 12 |
| PTCS 21 | T0 | 3 | 2 | 2 | 0 | 1 | 3 | 0 | 2 | 3 | 3 | 2 | 0 | 2 | 0 | 0 | 23 |
| PTCS 22 | T0 | 2 | 2 | 1 | 0 | 0 | 4 | 3 | 1 | 4 | 1 | 2 | 0 | 2 | 0 | 0 | 22 |
| PTCS 1 | T1 | 2 | 2 | 1 | 0 | 0 | 3 | 0 | 0 | 2 | 1 | 2 | 0 | 1 | 0 | 0 | 14 |
| PTCS 2 | T1 | 2 | 2 | 1 | 0 | 1 | 2 | 0 | 0 | 2 | 1 | 1 | 0 | 2 | 0 | 0 | 14 |
| PTCS 3 | T1 | 0 | 0 | 0 | 0 | 0 | 0 | 0 | 0 | 0 | 1 | 1 | 0 | 1 | 0 | 0 | 3 |
| PTCS 4 | T1 | 0 | 1 | 1 | 0 | 0 | 1 | 0 | 0 | 1 | 0 | 0 | 0 | 0 | 0 | 0 | 4 |
| PTCS 5 | T1 | 2 | 2 | 1 | 0 | 0 | 1 | 0 | 0 | 2 | 0 | 0 | 0 | 0 | 0 | 0 | 8 |
| PTCS 6 | T1 | 0 | 0 | 0 | 0 | 0 | 2 | 0 | 0 | 2 | 1 | 1 | 0 | 2 | 0 | 0 | 8 |
| PTCS 7 | T1 | 0 | 2 | 2 | 0 | 0 | 3 | 0 | 0 | 2 | 0 | 0 | 0 | 0 | 0 | 0 | 9 |
| PTCS 8 | T1 | 1 | 1 | 1 | 0 | 0 | 2 | 0 | 0 | 2 | 0 | 0 | 0 | 0 | 0 | 0 | 7 |
| PTCS 9 | T1 | 2 | 1 | 1 | 0 | 1 | 1 | 0 | 0 | 2 | 1 | 1 | 0 | 1 | 0 | 0 | 11 |
| PTCS 10 | T1 | 1 | 1 | 1 | 0 | 0 | 1 | 0 | 0 | 1 | 2 | 2 | 0 | 1 | 0 | 0 | 10 |
| PTCS 11 | T1 | 1 | 1 | 1 | 0 | 0 | 0 | 0 | 0 | 0 | 1 | 2 | 0 | 2 | 0 | 0 | 8 |
| PTCS 12 | T1 | 0 | 1 | 1 | 1 | 1 | 1 | 0 | 0 | 1 | 1 | 1 | 0 | 1 | 0 | 0 | 9 |
| PTCS 13 | T1 | 2 | 1 | 2 | 0 | 0 | 2 | 0 | 0 | 1 | 0 | 0 | 0 | 0 | 0 | 0 | 8 |
| PTCS 14 | T1 | 0 | 0 | 0 | 0 | 0 | 1 | 0 | 0 | 0 | 1 | 1 | 0 | 1 | 0 | 0 | 4 |
| PTCS 15 | T1 | 0 | 1 | 1 | 0 | 0 | 0 | 0 | 0 | 0 | 1 | 1 | 0 | 2 | 0 | 0 | 6 |
| PTCS 16 | T1 | 1 | 0 | 1 | 0 | 0 | 1 | 0 | 0 | 1 | 1 | 1 | 0 | 1 | 0 | 0 | 7 |
| PTCS 17 | T1 | 0 | 0 | 1 | 0 | 0 | 0 | 0 | 0 | 0 | 1 | 2 | 0 | 2 | 0 | 0 | 6 |
| PTCS 18 | T1 | 0 | 1 | 1 | 0 | 0 | 2 | 0 | 0 | 2 | 1 | 1 | 0 | 1 | 0 | 0 | 9 |
| PTCS 19 | T1 | 2 | 2 | 2 | 0 | 0 | 3 | 0 | 0 | 1 | 0 | 0 | 0 | 0 | 0 | 0 | 10 |
| PTCS 20 | T1 | 1 | 1 | 1 | 0 | 1 | 1 | 0 | 0 | 1 | 0 | 0 | 0 | 0 | 0 | 0 | 6 |
| PTCS 21 | T1 | 3 | 2 | 2 | 0 | 1 | 2 | 0 | 1 | 3 | 3 | 2 | 0 | 2 | 0 | 0 | 21 |
| PTCS 22 | T1 | 0 | 0 | 0 | 0 | 0 | 1 | 0 | 0 | 1 | 1 | 1 | 0 | 1 | 0 | 0 | 5 |

**Supplementary Table 4.** *Grand-Total EEG total score and subscores for each patient across time points (T0 and T1).*

*BA: background activity; EA: epileptiform abnormalities; PTCS: post-traumatic confusional state; SWA: slow wave activity; TBI: traumatic brain injury.*
