## Supplementary Table 5 for "EEG correlates of confusional state after traumatic brain injury"

| Drug Class |  | Wilcoxon rank sum test with continuity correction | p-value |
| --- | --- | --- | --- |
| Comparison: TBI control (T0) vs PTA/PTCS (T0) | | | |
| Sedatives-Hypnotics (Diazepam equivalent, mg) |  | 254 | 0.349 |
| Antipsychotics (Chlorpromazine equivalent, mg) |  | 206 | 0.726 |
| Antiepileptics (DDD, %) |  | 236 | 0.634 |
|  |  | **Wilcoxon signed rank test with continuity correction** | **p-value** |
| Comparison: PTCS (T0) vs PTA/PTCS (T1) | | | |
| Sedatives-Hypnotics (Diazepam equivalent, mg) |  | 25 | 0.838 |
| Antipsychotics (Chlorpromazine equivalent, mg) |  | 44 | 0.944 |
| Antiepileptics (DDD, %) |  | 17 | 0.308 |

**Supplementary Table 5.** *Comparison of drugs across clinical groups and timepoints. This table presents the statistical comparisons of drug use between TBI controls (T0) and patients with PTA/PTCS (T0), as well as longitudinal changes in drug use within PTA/PTCS between T0 and T1. Drug classes include sedatives-hypnotics (expressed as diazepam equivalents), antipsychotics (expressed as chlorpromazine equivalents), and antiepileptics (expressed as cumulative percentage of defined daily dose for each drug). The Wilcoxon rank-sum test found no significant differences in drug use between TBI controls and patients with PTA/PTCS at T0 (upper section), while the Wilcoxon signed-rank test showed no significant changes in PTCS patients over time (lower section).*

*DDD: defined daily dose; PTA/PTCS: post-traumatic amnesia/post-traumatic confusional state; TBI: traumatic brain injury.*
