## Supplementary figures and images for "EEG correlates of confusional state after traumatic brain injury"

### Supplementary Fig.1

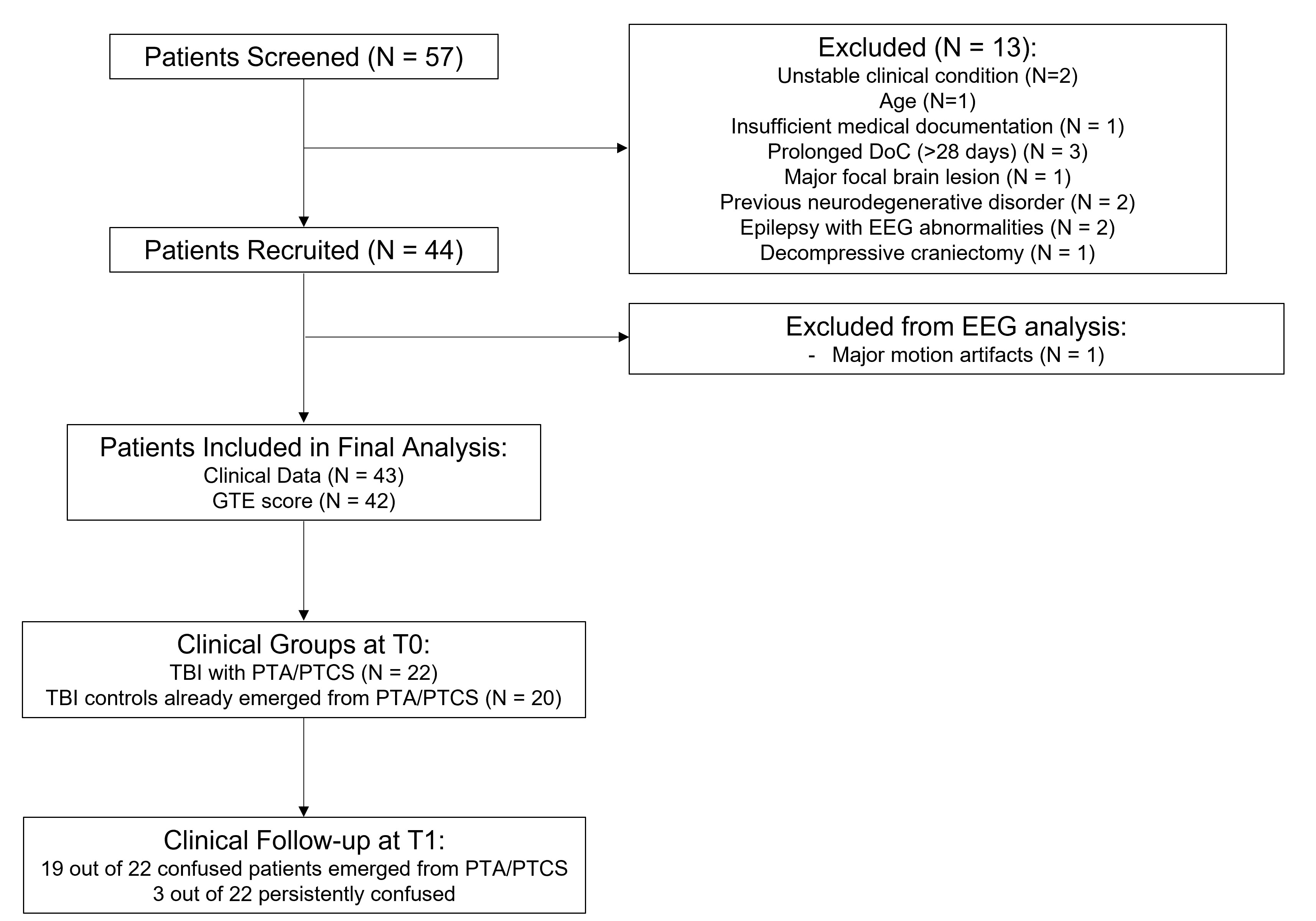
